# Reconstructing whole-brain structure and dynamics using imaging data and personalized modeling

**DOI:** 10.1101/2025.01.06.24319726

**Authors:** M. Fabbrizzi, L.G. Amato, L. Martinelli, J. Carpaneto, E. Bartolini, S. Calderoni, A. Retico, A. A. Vergani, A. Mazzoni

## Abstract

Brain structure plays a pivotal role in shaping neural dynamics. Current models lack the anatomical and functional resolution needed to accurately capture both structural and dynamical features of the human brain. Here, we introduce the FEDE (high FidElity Digital brain modEl) pipeline, generating anatomically accurate brain digital twins from imaging data. Using advanced techniques of anatomical tissue segmentation and finite-element analysis, FEDE reconstructs brain structure with high spatial resolution, while also replicating whole-brain neural activity. We demonstrated its application by creating the first brain digital twin of a toddler with autism spectrum disorder (ASD). Through parameter optimization, FEDE replicated both time-frequency and spatial features of recorded neural activity. Notably, FEDE predicted patient-specific aberrant values of excitation to inhibition ratio, coherently with ASD pathophysiology. FEDE represents a significant leap forward in brain modeling, paving the way for more effective applications of digital twin in experimental and clinical settings.

## Introduction

Computational brain models are widely utilized to investigate the structural underpinning of brain activity. Bioinformatics platforms such as The Virtual Brain (TVB)^1,2^, have emerged, enabling high-fidelity simulations of brain activity. Their use ranges from the identification of structural phenomena shaping brain dynamics in physiological conditions^3–5^, to clinical applications such as disease diagnosis^6^ and testing of therapeutic approaches^7,8^. However, the significant simplifications required to simulate and analyze virtual brain data efficiently have limited current models to replicating neural activity dynamics without achieving full anatomical accuracy.

Over the past years, several advanced MRI preprocessing and postprocessing techniques have been developed, enabling accurate studies of brain structure and functions in patients^9–14^. These advancements include methods to quantify voxel-wise myelination levels^11,15^, which are crucial for determining conduction velocity between brain regions^16^. Moreover, recent techniques take into account detailed anatomical properties, such as tissue anisotropy^17,18^, which significantly influence how brain activity is generated and propagated across brain tissues^19^.

Despite these advancements, there is currently no pipeline that successfully combines imaging analytic tools and computational models into a unified framework. Each technique requires specific software and packages (see Methods), resulting in a fragmented approach that limits integration. As a result, no existing model can achieve both an anatomically accurate reconstruction of the human brain and a high-fidelity replication of neural activity. This limits the potential to produce effective digital twins, i.e. virtual models that replicate the physiological and anatomical characteristics of the brain structure, capturing brain dynamics with the fidelity needed for comprehensive studies^20,21^.

To address these challenges, we present FEDE (high FidElity Digital brain modEl), a pipeline incorporating state-of-the-art imaging analysis tools and computational techniques to reconstruct brain anatomy and replicate individual whole-brain activity with high precision. FEDE incorporates advanced software to reconstruct detailed structural features of the human brain, including local cortical connections, voxel-wise myelination levels and tissue-specific anisotropy and conductance properties, shaping neural activity generation from cortical sources.

We demonstrated FEDE’s application by creating the high-fidelity digital brain twin model of a toddler (between 1y and 3y old) with autism spectrum disorder (ASD). Developing a digital twin of a toddler presents unique challenges, such as the incomplete myelination of frontal lobes^16^ and skull development, requiring tailored pipelines for young brain MRI analysis. Additional difficulties arise from the unclear etiology of ASD and the need to capture its multi-scale anomalies, ranging from synaptic dysfunction to whole-brain connectivity, without relying on predefined assumptions. ASD is a complex neurodevelopmental condition characterized by significant variability in its presentation and severity^22,23^. Several abnormalities in brain structure and function have been reported in ASD, including synaptic imbalance^24,25^, global and local connectivity alterations^26,27^, and abnormal brain growth^28,29^. Particularly, ASD is well documented to impair local connectivity within the brain^26,30^, which is crucial for cognitive and sensory processes^27^. Traditional models of brain activity often overlook the importance of this local connectivity, focusing instead on global measures of connectivity^31,32^ or synaptic transmission^27,33,34^. Inter-subjective heterogeneity makes ASD an ideal candidate for the use of digital twins^20^, as both structural and dynamical alterations contribute to the condition’s pathophysiology.

FEDE replicated whole-brain neural activity of the patient (measured via EEG recordings) with high precision, accurately reproducing both time-frequency domain features, such as power spectral density (PSD), and spatial features, such as functional connectivity. Moreover, it identified aberrant values of excitation to inhibition ratio as the structural culprit of the experimental neural activity dynamics, consistent with ASD pathophysiology.

## Results

In this study, we present FEDE: a high-precision pipeline for the creation of personalized digital twin model leveraging comprehensive MRI data (Figure 1A), including T1-weighted (T1-w), T2-weighted (T2-w), and Diffusion-Weighted Imaging (DWI). Briefly, the model extracts MRI features (Figure 1B and Methods) to reconstruct brain properties (Figure 1C left and Methods), which are then used to simulate EEG signals (Figure 1C right and Methods). These simulated EEG signals can capture spatiotemporal organization of recorded EEG signals of the reconstructed brain (Figure 1E) and from this infer properties of the underlying structure and function (Figure 1E). The pipeline was tested on a toddler (aged between 1y and 3y) with autism spectrum disorder (ASD), to reconstruct an anatomically accurate digital twin replica of the patient’s brain and to replicate with high fidelity the experimental EEG recordings.

**Figure 1:**
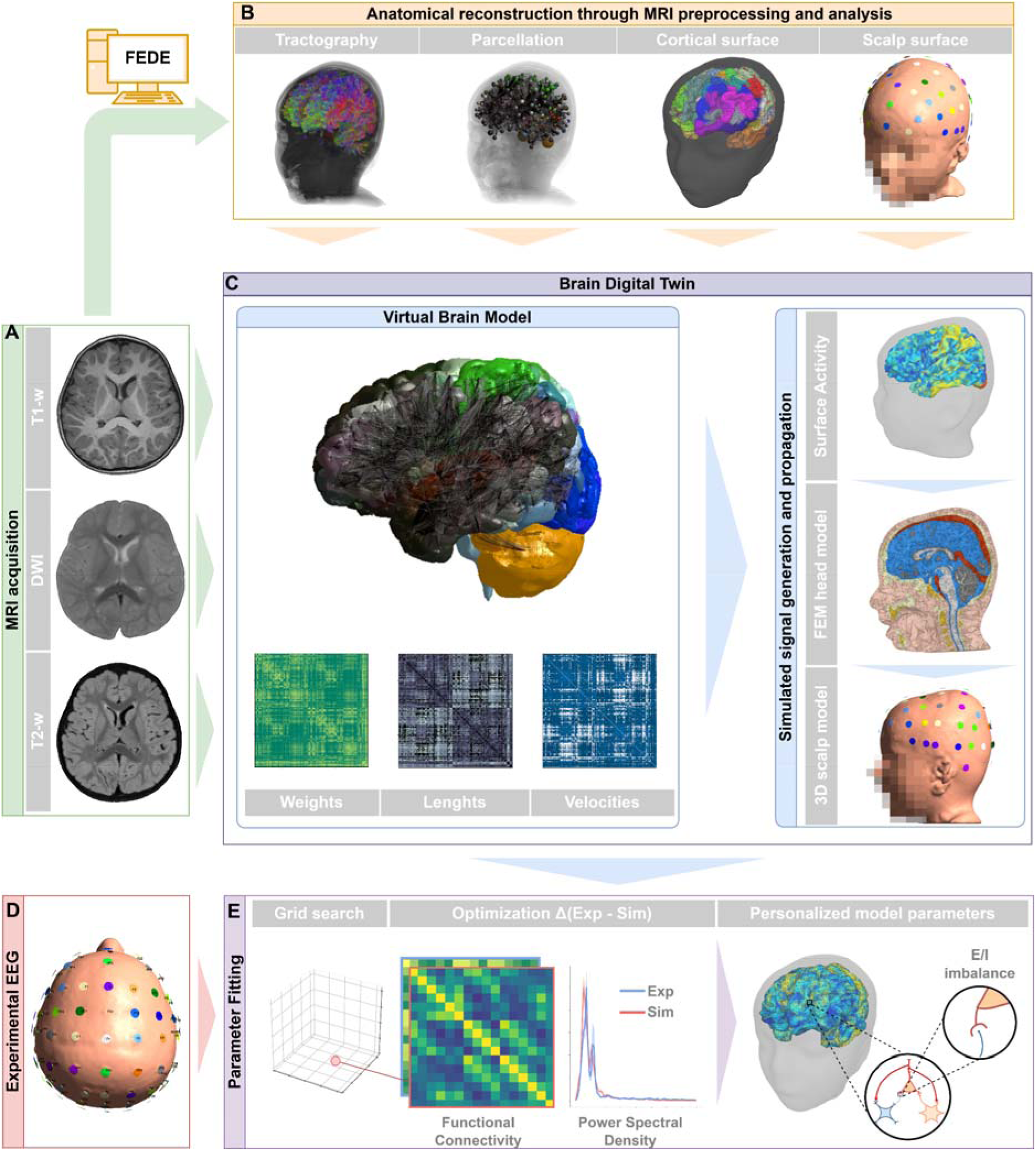
The FEDE (high FidElity Digital brain modEl) pipeline: (A): MRI recordings, including (from top to bottom) T1-w, DWI and T2-w, were used to generate a 3D replica of the patient brain. **(B):** MRI processing steps: from left to right anatomical constrained tractography analysis, brain regions parcellation and segmentation according to the HCPMMP1 atlas. Right end: reconstructed 3D model of the patient’s head, including a high-density mesh of the cortical surface and a scalp model with EEG electrode positions. **(C):** Bottom: the parcellation of brain areas allowed to compute connective weights, distances and conduction velocity maps for the whole brain structure, which were integrated in a virtual brain model (top, see Methods). Top right: Neural mass models were used to compute neural activity on the high-density cortical surface. Middle right: The activity was then projected to the scalp of the patient with an anatomically-accurate lead-field matrix, leveraging a FEM model of the patient’s head and including tissue anisotropy of brain tissues. Bottom right: This allowed for the computation of EEG signals from simulated brain activity. **(D):** EEG recordings were acquired during resting-state and EEG features such as power spectral distribution and functional connectivity are extracted. **(E):** Optimal fit between experimental and simulated EEG led to the identification of structural parameters underlying patient’s condition. Parameter inversion, based on a grid search on candidate parameters, allowed to infer personalized parameters that replicated at best the experimental EEG recordings of the patient.

### Personalized conduction velocity map is needed for accurate reconstruction of neural connections

The FEDE pipeline comprises the individual reconstruction of neural connections from MRI data. Brain regions were parcellated according to the HCPMMP1 atlas^35^, encompassing 379 cortical and subcortical regions. Connections between regions were computed from DWI data (Supplementary Figure S1) and tract lengths (the average length of white matter fiber. bundles connecting two regions) were determined from tractography DWI analysis. Crucially, alongside standard tractography, FEDE incorporates the computation of conduction velocities from brain areas, which considers voxel-wise levels of myelination and their role in determining velocity of signal transmission across brain areas.

In FEDE, voxel-wise levels of myelinization are first computed from T1-w and T2-w imaging combined with apparent fiber density (obtained from DWI^36^, see Methods) using Equation 1. Conduction velocity map is then computed from Equation 2, enabling a detailed analysis of signal transmission properties across the brain (Fig 2A). The conduction velocity map offers an improved method for understanding how signals propagate through different brain regions by considering the anatomical and microstructural properties of brain tissues, primarily myelination of white fibers. Notably, a lower conduction velocity can be observed in the frontal lobe (Fig 2A panel iv) due to the young age of the subject and the myelination process not completed. This introduces topographic differences in conduction velocities that would be overlooked using standard methodologies.

**Figure 2:**
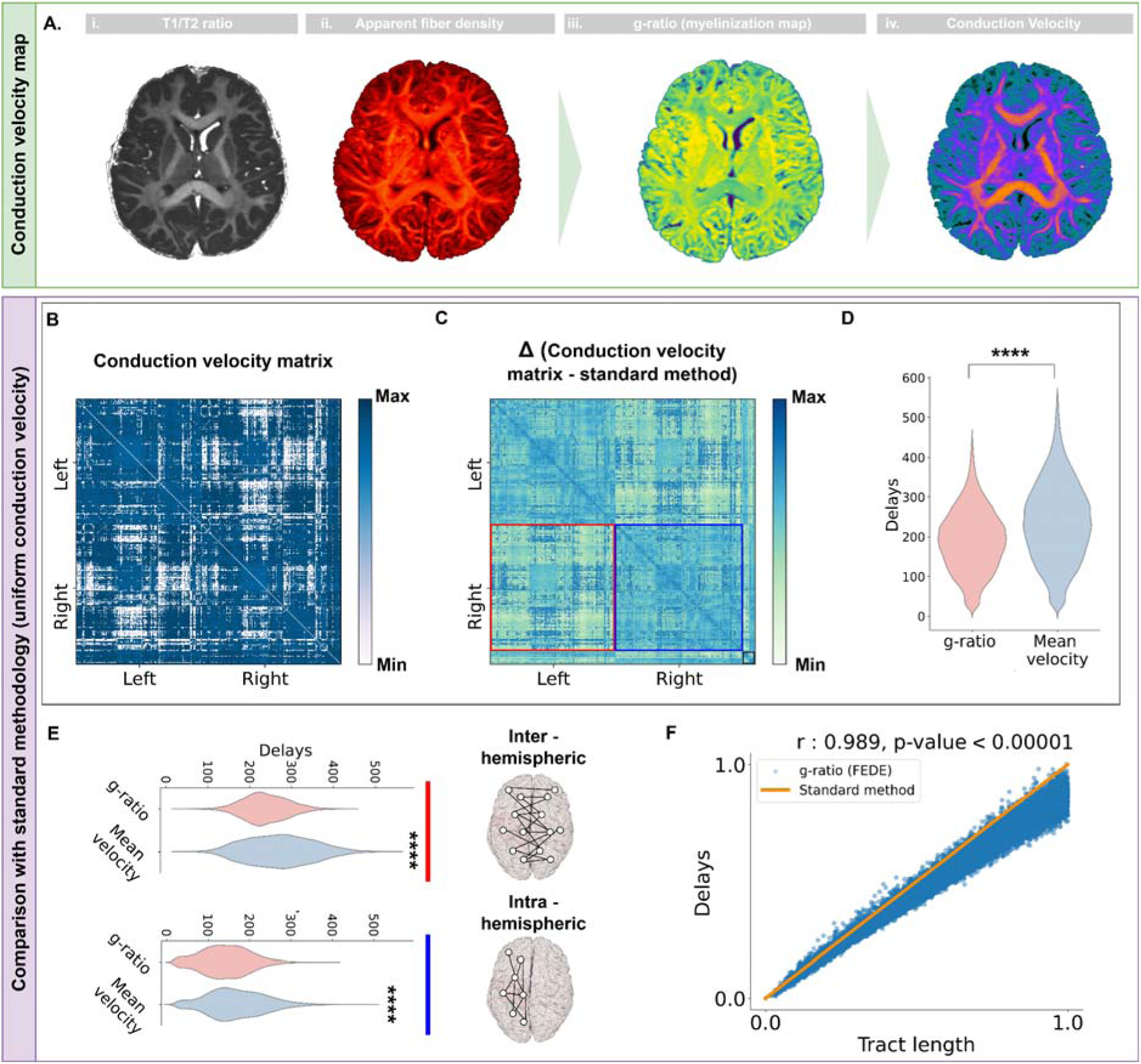
Delays in neural activity propagation are significantly overestimated by standard methodologies. **(A)**: Conduction velocity map is computed from the ratio between T1-w and T2-w (i), combined with apparent fiber density (ii) obtained from DWI (see Methods). It allows for the computation of axonal myelin levels, defined as the g-ratio (iii), which leads to a map representing voxel-wise levels of conduction velocity of brain signals (iv). Orange indicates high values, while low values of conduction velocities are indicated in blue. **(B)**: Conduction velocity matrix *CV_a,b_* whose entries encode the conduction velocity from region *a* to region *b* **(C)**: Difference between delays computed dividing the tract lengths matrix by the *CV* matrix (*TL/CV*) and delays computed with standard methodology (i.e. from the tract lengths matrix and a mean conduction velocity value). Red (blue) square represents inter-hemispheric (intra-hemispheric) connections, while black box represents subcortical regions. **(D)**: Violin plot of delay values computed with the FEDE g-ratio method (red) and standard method (cyan). Delay values computed from a mean conduction velocity value tend to overestimate with high significance actual delay values computed from MRI analysis. **(E)**: Violin plot of delay values computed with the two methods, separately for interhemispheric connections (top) and intra-hemispheric connections (bottom). Inter-hemispheric delays are significantly overestimated with standard methodology (see main text). **(F)**: Scatter plot of tract length values and delays computed using the *CV* matrix obtained with FEDE. Orange line represents the expected delays according to standard methodology. Significance notation: **** stands for p<0.00001.

The model was then used to combine the voxel-wise map with the selected gray matter parcellation. This led to the computation of the patient-specific conduction velocity matrix *CV_a,b_* Each entry of the matrix encodes the conduction velocity between region *a* and region *b* in the adopted parcellation (Fig 2B). This matrix allows for the calculation of delays, dividing the tract lengths matrix by the conduction velocity matrix (*TL/CV*).

We compared the delays computed using FEDE with those derived from the standard methodology, in which delays are simply proportional to the tract lengths matrix. Significant discrepancies were found between the two methods (Fig 2C). Delays calculated with FEDE were significantly lower with respect to those computed with standard methodology (ratio of 0.79±0.05, Mann-Whitney test, U=1.3e+11, p<0.00001), suggesting that delays computed using standard methods tend to overestimate the actual values.

The overestimation of delay times by standard methodology was further analyzed by examining inter-hemispheric and intra-hemispheric connections separately (Fig 2E). For inter-hemispheric connections, delays were significantly larger when using the standard methodology (delays computed with FEDE were 0.72±0.04 of those computed with standard methodology, U=5e+9, p<0.00001). Intra-hemispheric connections also showed statistically significant differences, though less pronounced compared to inter-hemispheric connections (ratio was 0.87±0.05, U=8e+9, p<0.00001).

The overestimation of delay times with standard methodology was confirmed by a pairwise comparison between delay times computed with FEDE and with standard methodology (i.e., from the length of connective tracts between regions). Delay times computed with FEDE strongly correlated with tract lengths (r=0.989, p<0.00001, Fig 2F), with relevant differences from the exact proportionality postulated by standard methodology, (orange line in Fig 2F). Delay times computed with FEDE were found to be smaller than values expected from the standard methodology (dots below the orange line in Fig 2F) in 96% of the cases. The significance of the pairwise differences in delay times was confirmed by a ^2^ test (test=7.8e+5, p<0.00001). A distribution analysis between FEDE and standard methodology also confirmed statistically significant differences (Kolmogorov-Smirnov test=0.086, p<0.00001). This consistent overestimation introduces a significant bias in models built on standard delay values, potentially affecting the accuracy of any subsequent analysis on functional connections and brain activity.

### Anatomically-accurate lead field matrix allows to precisely deduce cortical sources of simulated EEG signals

To achieve a high-fidelity replication of individual neural activity, we computed an anatomically-accurate forward-solution utilizing personalized tissue reconstruction from MRI data (Fig 3A). In FEDE, neural activity is simulated using the Jansen-Rit neural mass model^37^ on a high density cortical mesh of 20.484 vertices. The simulated cortical activity is then projected onto the scalp surface via a lead-field matrix (LFM) meticulously derived from the patient’s anatomical data (see below and Methods). This matrix ensures that the projection of neural signals to the EEG electrodes accurately reflects the patient’s unique brain anatomy^38^.

**Figure 3:**
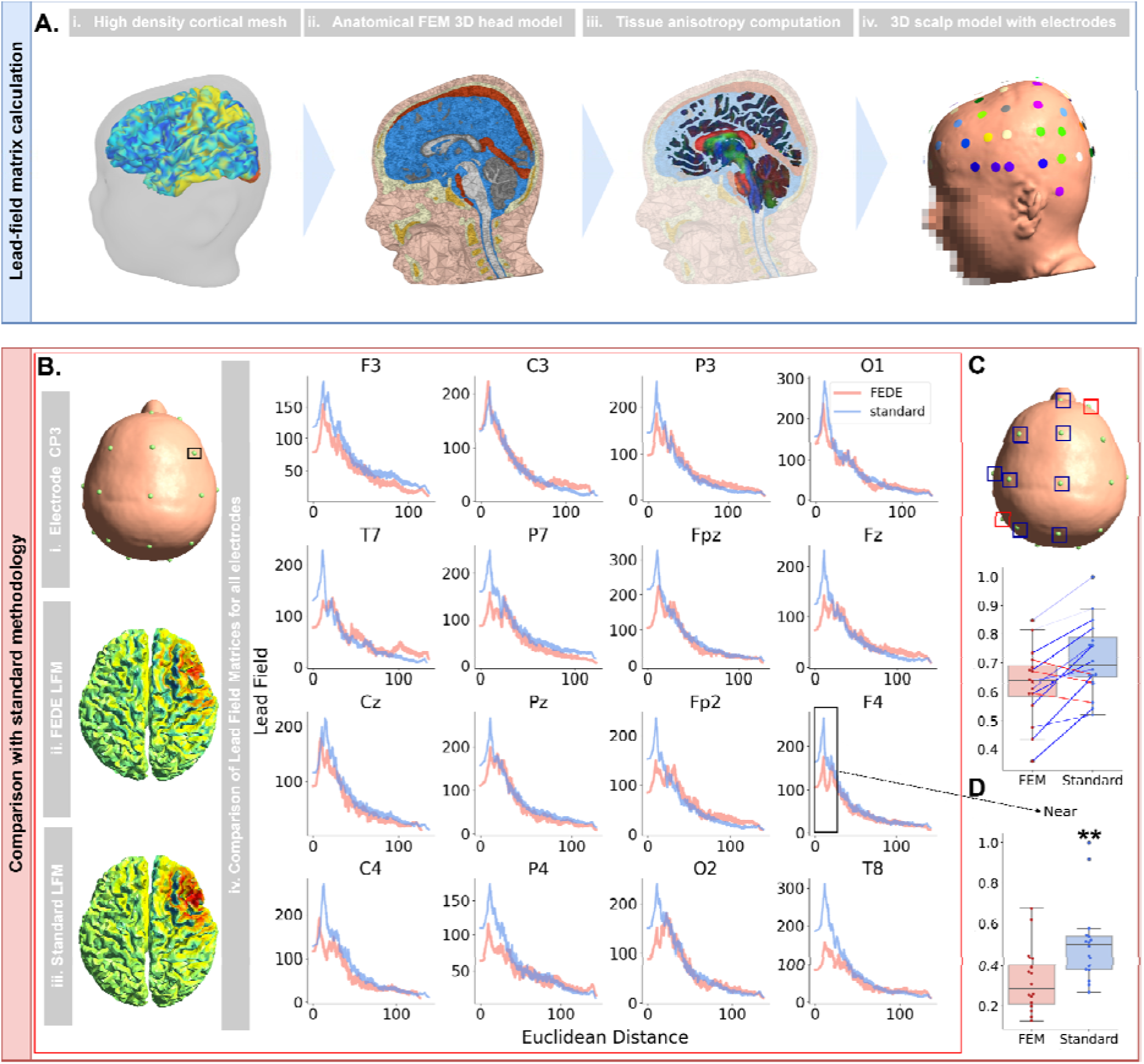
Anatomically-accurate forward-solution with personalized tissue reconstruction identifies correct cortical sources of each EEG channel. (A): From left to right: (i) Cortical activity computed on the high-density cortical mesh; (ii) anatomical tissues, each with its own conductance value, were reconstructed with SIMNIBS software, creating a 3D FEM model of the patient’s head, which was combined with tissue anisotropy computed from DWI data (iii). This allowed for the precise propagation of the electric field onto the scalp surface, and thus to the electrode grid (iv). This procedure results in the reconstruction of the cortical contribution to the activity of each electrode. **(B):** Different LFMs for the F4 electrode (i) plotted as an example, with FEDE model LFM (ii) and standard method LFM (iii); In each EEG channel, LFM contribution as a function of the distance to the electrode (iv) presented strong differences between FEDE model (salmon) and standard method (light blue). Only channels selected for experimental analysis are shown. **(C):** Scalp view and boxplot of LFM values according to both standard methodology and FEDE. Statistical tests revealed that standard method resulted in an overestimation of LFM values in most EEG channels (indicated by blue color in the boxplot and in the scalp plot), while it resulted in an underestimation only in two channels (indicated by red color in the boxplot and in the scalp plot). Only channels selected for experimental analysis are shown, and channels with significant differences across methods were highlighted. Whiskers represent interquartile ranges. **(D):** Standard methodology significantly overestimates contributions from near vertices. Near vertices are defined (for each channel) as the 20% of closest vertices; in panel (B) is reported the example of LFM values of near vertices for the F4 electrode. Notation is the same as in (C). Significance notation: ** stands for p<0.01, Bonferroni correction.

To generate an anatomically-accurate LFM, FEDE constructs a finite element model (FEM) of anatomical tissues from MRI data using the SIMNIBS software (see Methods). This approach allows the LFM computation to account for ten distinct tissue types (plus two for electrodes and saline solution used on the patient scalp), each with its specific conductivity and anisotropic properties (see Supplementary Fig. S2).

We compared this anatomically-accurate LFM to one derived from standard boundary element methods (see Methods). For each electrode, we calculated the cortical contributions to the LF matrices, organizing cortical vertices by their distance from the electrode (Fig 3B). A Kolmogorov-Smirnov test revealed significant differences in the distributions of LFM values between the two matrices for all electrodes (p<0.00001). We analyzed LFM values channel-wise, to assess if standard methodology introduced a significant bias in determining LFM contributions for different electrodes. Significant differences were found in 11/16 electrodes. The standard method overestimated contributions for 13/16 electrodes, with significant overestimation (p<0.05) in 9 electrodes. For the remaining 3 electrodes, standard methodology caused an underestimation though this difference was statistically significant only in two of them. Notably, the bias introduced by standard methodology was strongly lateralized, with 10 of the 11 electrodes that presented statistically relevant differences located in the central or left portion of the scalp (Fig 3C).

Additionally, we conducted a separate analysis for vertices near to the electrode position (defined as the 20% closest vertices for each electrode), to assess whether standard methodology overestimated their LFM contributions. Quantitative analysis confirmed this hypothesis, with the standard method significantly overestimating contributions from near vertices (0.32±0.08 for FEM vs. 0.50±0.10 for standard method, Fig 3D), with a strong statistical significance (U=57, p=0.008). On the contrary, differences between the two methods were non-significant for vertices far from the electrodes (defined as the top 20% farthest vertices for each electrode, U=113, p=0.58). The interaction between the distance from the electrode and the differences in LFM contribution between standard method and FEDE was confirmed by a two-way ANOVA (f=3.49, p=0.066).

In summary, the standard method introduced substantial bias in estimating cortical contributions to EEG signals, particularly by underestimating the effects of volume conduction and overestimating cortical source specificity.

### FEDE replicates accurately the spatiotemporal structure of experimental EEG recordings

Leveraging its anatomically accurate 3D cortex and head model, FEDE allowed to simulate individual brain activity with high precision. We validated the model by reproducing the previously acquired experimental recordings of the toddler with ASD (see Methods).

We determined the combination of model parameters leading to the optimal replicated the experimental EEG signals. We first analyzed the PSD of the digital twin model, finding high resemblance to the experimental one (Fig 4A). The analysis of different EEG frequency bands (Fig 4B) further demonstrated that the relative power of the digital twin’s PSD correctly replicated the theta and delta dominant rhythm observed in the experimental recording supporting the model’s reliability in simulating distinct oscillatory behaviors. Linear regression between simulated and experimental PSD values confirmed the high accuracy of the digital twin, with a correlation value r=0.92, r^2^=0.84 p<0.00001.

**Figure 4:**
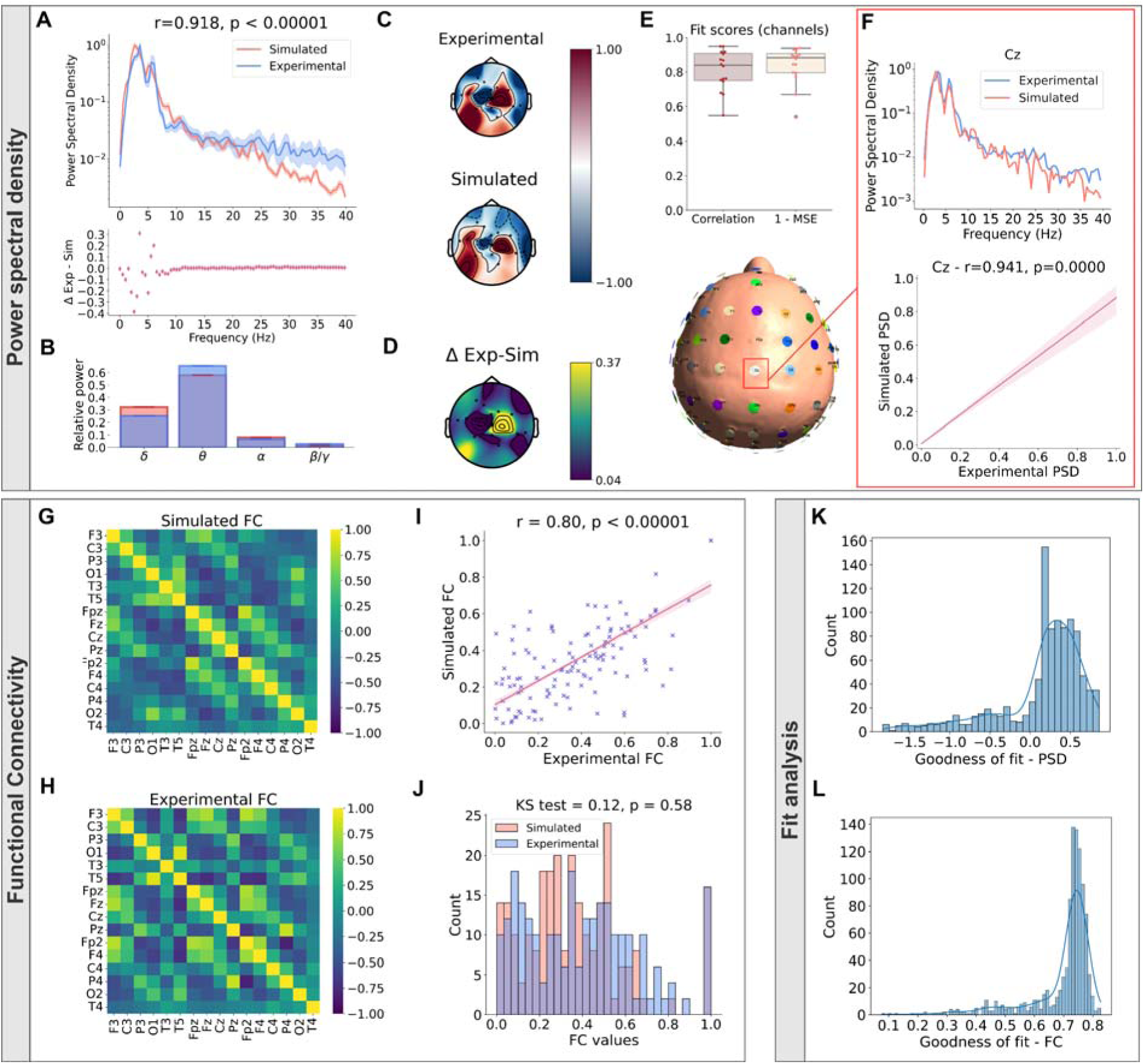
FEDE spatiotemporal accuracy in replicating experimental EEG signals (A): Patient’s average PSD (top plot, light blue) superimposed to the average PSD of the digital twin (salmon). Bottom inset shows residual plot. The r-value between simulated and experimental data was of 0.918. **(B):** Relative power in different EEG bands for the patients and the digital twin. Notation is the same as in (A). **(C):** Topographic plot of scalp EEG activity in the patient and in the digital twin, indicating that the model can reproduce the non-trivial topographies encountered in experimental recordings. Signals were normalized between-1 and 1. **(D):** Topographic plot of the (absolute) difference between experimental and simulated EEG activity reported in (C). **(E):** Fit scores of the digital twin, reporting correlation values and accuracy (measured as 1 – mean squared error) between simulated and experimental recordings. Each point represents a different EEG channel. Note how correlation was > 0.6 for all channels. **(F):** Cz channel is reported as an example, with PSD (top plot) and regression line between experimental and simulated PSD values (bottom plot). Notation is the same as in (A). **(G):** FC matrix computed from simulated recordings of the digital twin. **(H):** FC matrix computed from patient’s experimental recordings. **(I):** Linear regression between simulated and experimental FCs, with correlation value reported. The digital twin’s FC had a correlation of 0.80 with the experimental one. **(J):** Histogram of FC values for the patient and the digital twin. Kolmogorov-Smirnov test highlighted no significant differences between the two distributions. Notation is the same as in (A). **(K):** PSD Goodness of fit analysis for tested combination of model parameters (only values greater than 0 are reported). **(L):** FC Goodness of fit analysis for tested combination of model parameters (only values greater than 0 are reported). While most combinations of model parameters produced FCs similar to the experimental one, only few combinations allowed to replicate the PSD.

Additionally, the model reproduced the complex topography observed in experimental recording (Fig 4C), maintaining consistent correlations with experimental EEG across all channels (see Fig 4D for differences between the two). The average correlation between simulated and experimental single channel PSDs was r=0.81 ± 0.11, and a mean squared error analysis yielded a 1-MSE value of 0.84 ± 0.10 (Fig 4E). These results indicated that the FEDE not only captured individual channel characteristics but also preserved a high degree of accuracy in simulating spatial EEG patterns. Channel-wise comparisons between simulated and experimental PSDs can be found in Supplementary Fig. S3A (an example channel is found in Fig 4F). We also conducted a single channel analysis for each frequency band, to assess the ability of the digital twin in replicating the experimental relative powers in each band for each channel. Kolmogorov-Smirnov test conducted between simulated and experimental distributions of relative powers revealed no difference in the EEG frequency bands (Supplementary Table 2, distributions are found in Supplementary Figure S3B).

The digital twin also correctly replicated experimental FC. When comparing functional connectivity matrices derived from both simulated (Fig 4G) and experimental recordings (Fig 4H), the digital twin produced FCs with strong correlation to experimental data (Fig 4I, r = 0.80, *r*^2^=0.64, p < 0.00001). A Kolmogorov-Smirnov test comparing the distributions of FC values from the two data sets found no statistically significant difference (test = 0.13, p = 0.58), indicating that the model successfully replicated the complex patterns of brain connectivity observed in the experimental recordings (Fig 4J). A channel-by-channel comparison further corroborated the absence of relevant differences between simulated and experimental signals in FC values (Mann-Whitney U test, U=159.0, p=0.25, not shown). FC replication was also repeated with metrics less affected by volume conduction such as phase slope index^39^, obtaining similar results (r=0.53, p<0.00001).

Despite the generally good performance in replicating experimental EEG features, only few parameter combinations yielded precise replication of experimental PSD (Fig 4K), while many combinations allowed for an accurate replication of experimental FC (Fig 4L). This finding may be due to the influence of anatomical connections in shaping FC patterns (see Discussion) which highlights once more the importance of the digital twin model used as a scaffold to simulate brain activity.

We assessed the ability of the model constructed with FEDE to replicate EEG signals, by comparing it to a model constructed using a methodology not including the advancements introduced with FEDE. Specifically, we calculated the LFM using a standard 3-layer simplified Boundary Element Method (BEM) model of the head without anisotropy and with region-based simulations (see Methods). The structural connectivity matrix implemented in the standard model is the one obtained with the FEDE preprocessing pipeline, including anatomical constrained and probabilistic tractography. We conducted another parameter search to identify the optimal parameter combination for this standard approach. In terms of functional connectivity, the standard approach resulted in reduced fidelity with respect to FEDE (Fig 5A), with the correlation coefficient declining from r=0.80 to r=0.75. Regarding PSD, results indicated a notable decrease in fidelity for simulated PSD when compared to the FEDE model; the correlation (r-value) between experimental and simulated PSDs dropped from 0.92 to 0.60. Furthermore, this method produced simulated signals with a different dominant frequency than the experimental PSD, and it lacked key features such as the secondary peak in the PSD (Fig 5B).

**Figure 5:**
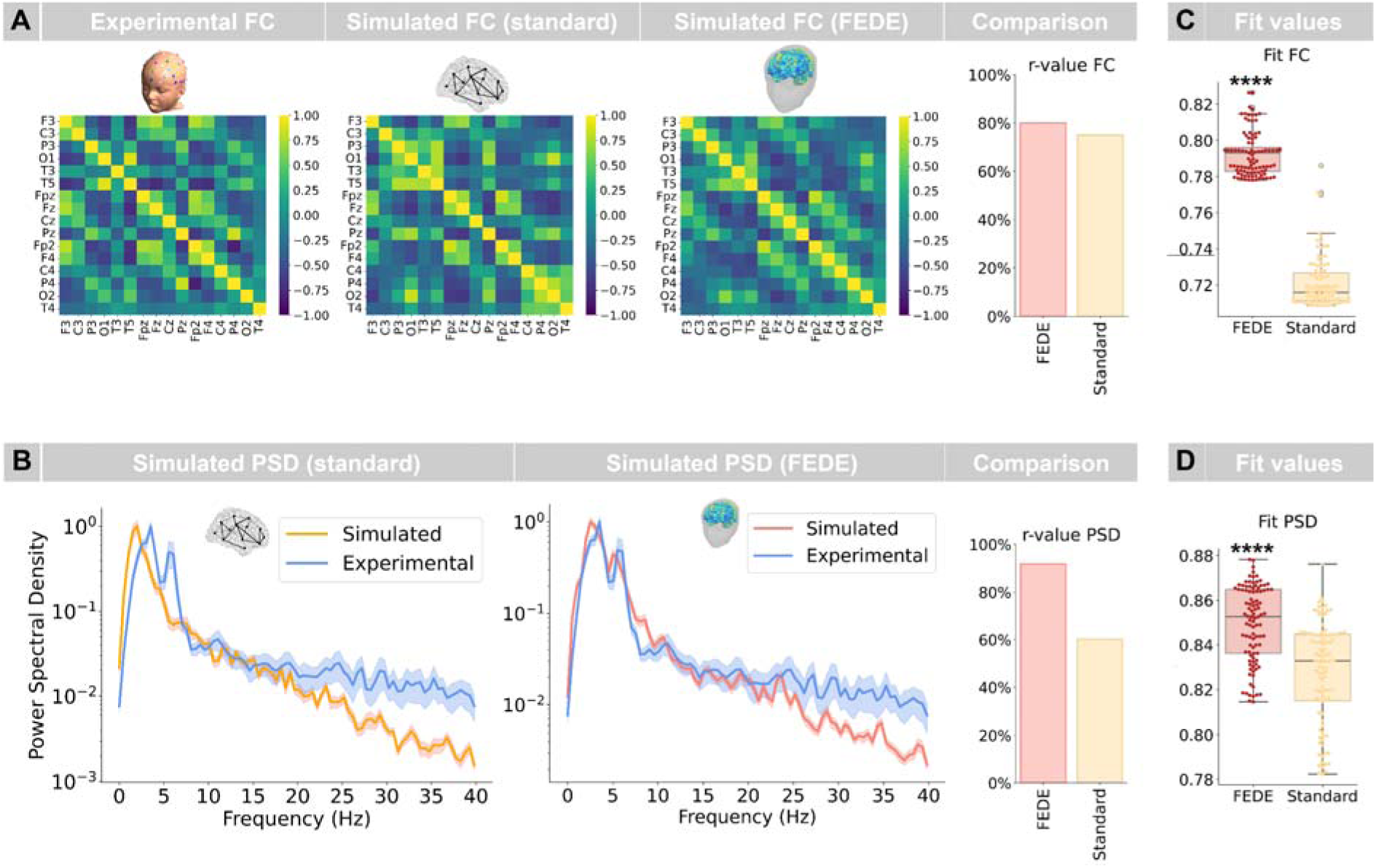
FEDE outperforms standard digital twin models in replicating experimental EEG features. Model parameters selected through parameter exploration were utilized in a region-based simulation, using as scaffold the 379-regions HCPMMP1 atlas employed for gray matter parcellation. No conduction velocity map was included, and both Lead-field matrix and 3D head model were reconstructed using standard methodology (see Methods). Results were compared with both the experimental values and with surface-based simulations. **(A):** FC matrices computed from standard digital twin showed different patterns with respect to those observed in experimental FC. For FC also, standard digital twin simulation presented a smaller Pearson r-value with experimental FC compared to the value obtained with FEDE. **(B):** PSD computed from region-based simulations present reduced similarity with experimental signals, presenting different dominant frequency and no second peak in low-alpha band. This caused the standard digital twin simulation to present a smaller Pearson r-value with experimental PSD compared to the value obtained with FEDE. **(C):** Fit values between experimental and simulated FC for 100 best standard and FEDE simulations. FEDE presented higher fit values in almost all simulations. **(D):** Fit values between experimental and simulated PSD for 100 best standard and FEDE simulations. FEDE presented higher fit values with high significance. Significance notation: **** stands for p<0.00001.

To evaluate the robustness of both methods, we analyzed the top 100 parameter combinations for each, comparing their fit functions with experimental FCs (Fig 5C) and PSDs (Fig 5D). In both cases, simulations generated by the FEDE model significantly outperformed those generated by the standard method (Mann-Whitney U test: U=9969, p<0.00001 for FC; U=7685, p<0.00001 for PSD).

Notably, the similarity between experimental and simulated FC remained high even in the simplified model, probably reflecting the high anatomical accuracy of the connectome reconstruction obtained with FEDE (considering that both models utilized the FEDE-derived connectome). However, the absence of anatomical details in the LFM and in the 3D head replica of the standard model significantly decreased its ability to capture finer details of the experimental PSD such as the second peak at low alpha frequency.

### Hierarchical parameter fitting to reconstruct personalized structural parameters of the patient’s brain

To determine the best combination of model parameter that replicated at best experimental EEG signals (see Fig 4), we simulated EEG signals from various parameter combinations (see Methods). We then analyzed their performance in replicating key metrics such as power spectral density and functional connectivity. These simulated signals were compared to the actual EEG recording of the patient, using the hierarchical parameter fitting procedure implemented in FEDE, based on the loss functions reported in Equations 3-4 (see Methods).

In each step of the procedure (Fig 6A), FEDE operates a grid search over a range of candidate parameters, identifying the optimal parameter combination by minimizing the loss functions. In the first step, FEDE determines the optimal values of parameters describing connections between brain areas. These parameters include the global connectivity coupling, the area and the strength of the local connectivity gaussian kernel (Step 1 in Fig 6A), and the conduction velocity proportionality constant. The second step comprises the identification of parameters determining the frequency output of the Jansen-Rit model, including the post synaptic potentials, noise and time constants of excitatory and inhibitory subpopulations, as well as the mean input firing rate. The ratio between inhibitory and excitatory time constants allows to determine also the excitation to inhibition ratio (EI ratio) of the brain. The last step involves the tuning of Jansen-Rit parameters that determine finer effects on output activity, such as the total number of connections in the neural mass, and the magnitude of links between single subpopulations and the sigmoid term transforming membrane potential into firing rate^37^. At each step, the initial point of the grid is determined by the best match identified in the previous step.

**Figure 6:**
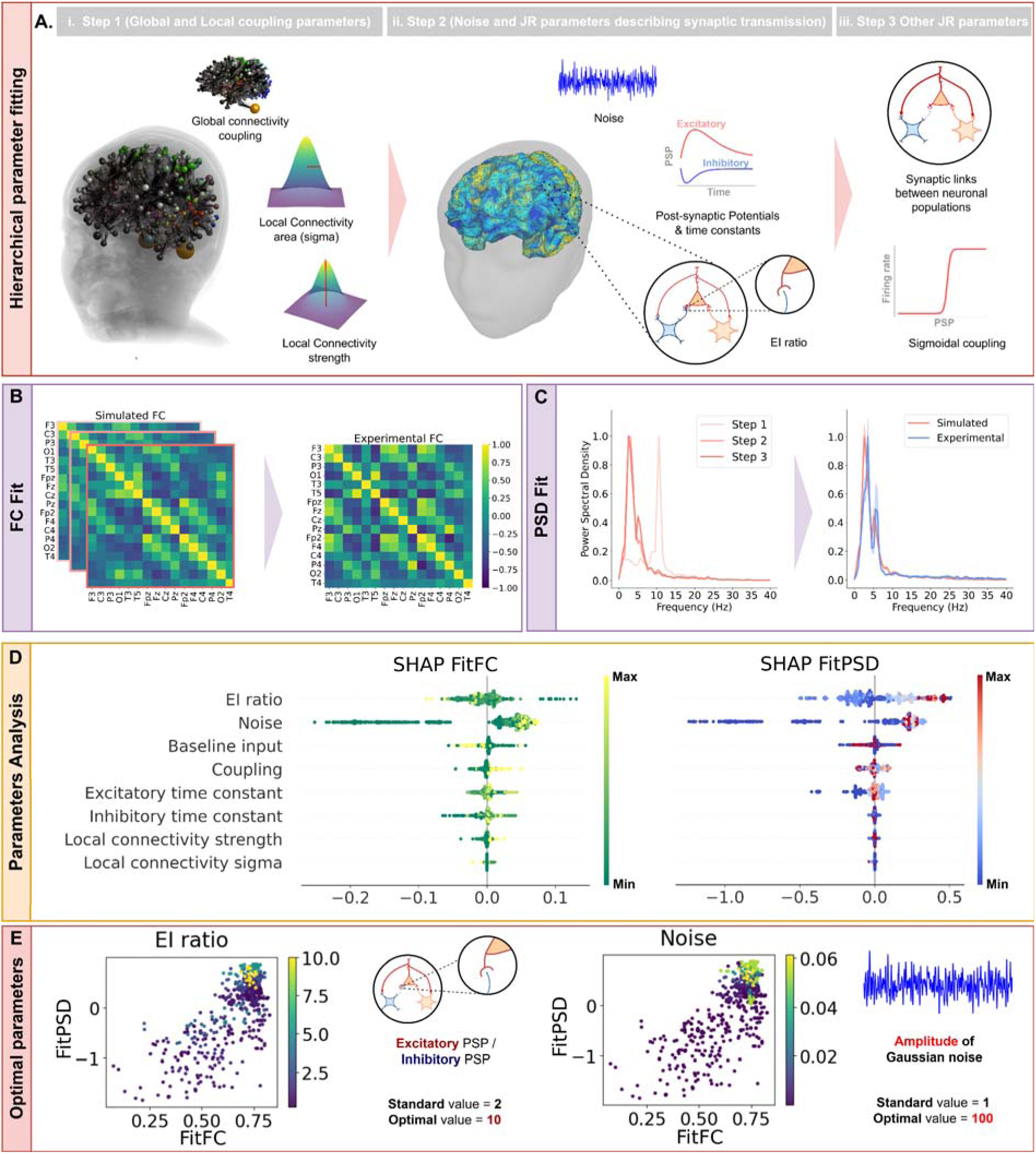
Hierarchical parameter fitting was employed to replicate EEG features and reconstruct underlying structural parameters. (A): The parameter fitting procedure followed three steps. At each step, the procedure identified a combination of model parameters, that was used as starting point for the next step of the parameter fitting procedure. The first step (i) identifies values of structural parameters of global and local connectivity, as well as velocity proportionality constant. The second step (ii) identifies Jansen-Rit parameters that dictate the frequency output of the model and mean input firing rate. The third step (iii) identifies Jansen-Rit parameters that govern finer features of the simulated output (see for example the second peak of the PSD). The last step focused on the sigmoid transforming membrane potential into firing rate. **(B-C):** At each step, the fidelity in replicating both FC and PSD increases, thanks to the identification of more precise parameters. **(D):** The parameter analysis module allows to investigate the importance of each parameter in correctly replicating experimental EEG activity, including statistical tools like SHAP analysis (in the labels Baseline input represents the total input to excitatory interneurons as sum of mean input firing rate, noise, global and local coupling outputs). **(E):** The pipeline allows to identify the structural parameters that more robustly determine EEG activity, that in our patient were aberrant values of the EI ratio and of background noise. FEDE includes tools for the visualization of fit values in relation to model parameters values. In the two scatter plots each dot is a different simulation, higher values of the corresponding parameter (respectively EI ratio and noise) are reported in yellow. See how higher values of both parameters allowed to obtain high fit values for both PSD and FC.

In our patient, this procedure allowed to increase at each step the precision of both simulated FC (Fig 6B) and PSD (Fig 6C). FEDE also comprises a module for the analysis of fitted parameters, allowing to identify the structural brain parameters playing the most relevant role in the agreement with experimental EEG activity of the patients (see Methods and Fig 6D). A SHAP analysis (see Methods) is implemented to quantitatively identify the model parameters that allow to replicate at best experimental EEG activity. The analysis revealed that only two parameters presented relatively high SHAP values, being EI ratio (0.0033 0.008) and noise (0.0041 0.024), while all other parameters presented values <0.00001.

Since each model parameter describes biophysical quantity closely related to microscopical mechanisms of neural connectivity and synaptic transmission, the optimal values identified by the procedure can be compared with physiological values to support diagnosis of pathological conditions. In our patient, this approach led to the identification of aberrant values of background noise (whose optimal value was 100 times greater than the standard one^2,37^, Fig 6E) and EI ratio (found to be five times greater than the standard healthy value^37^, see Fig 6E), which were essential in correctly replicating the experimental EEG activity. Aberrant values of both parameters are consistent with current hypotheses on ASD pathophysiology^24,25,40,41^.

## Discussion

The FEDE pipeline generated a high-fidelity digital twin of the brain of a with ASD and to replicate experimental EEG signal with unprecedented precision, while concomitantly identifying structural culprits of relevant EEG features. This was achieved by integrating state-of-the-art methods for MRI analysis, brain modelling and EEG simulation, and represents a substantial leap forward in the use of computational brain models in clinical settings, particularly in the context of personalized medicine.

The use of a detailed conduction velocity map derived from T1-w, T2-w, and DWI data provides a more accurate and nuanced understanding of neural signal transmission delays. It corrects the biases introduced by the traditional approach, ensuring that the physiological variability in conduction velocity across white matter fibers is properly accounted for. In Newman et al.^16^ a similar procedure based on the derivation of a conduction velocity map from MRI recordings highlighted significant differences between healthy and ASD subjects. Our approach allows not only to replicate the personalized determination of conduction velocity maps, but also to assess the role played by this quantity in shaping whole-brain activity.

A critical component of our methodology was the precise computation of the lead-field matrix, which defines the contribution of electrical activity from different cortical regions to the signals recorded by each EEG electrode. Our high-resolution cortical model allowed for an accurate calculation of this matrix, minimizing potential errors in source localization. This accuracy was reflected in the detailed comparison of the simulated and experimental EEG data, where the spatial correlations of neural activity (the FC matrix) closely replicated real-life recordings. In current computational brain models, the LFM is derived from an approximated boundary element method based on the compartments between brain, skull and scalp^7,8,42,43^. However, this approach neglects several aspects of brain anatomy, such as orientation of cortical sources and presence of several intermediate tissues between sources and electrodes with different conductivity values.

In the FEDE pipeline the EEG activity was computed from local sources on a high density (20,484 vertices) cortical mesh. In Wang et al.^8^ neural activity is simulated from a high density cortical mesh of more than 200,000 vertices. However, this procedure required to perform model inversion with a neural mass model-based simulation of 180 nodes, with parameters that were then translated to the surface simulation on the cortical mesh, in which neural activity was computed with a neural field model approach. In our study, we found a decrease in the fidelity of simulated neural signals (see Supplementary Fig S4 for an example). Our approach includes the parameter fitting procedure directly on the activity computed on the cortical mesh, enabling a more precise replication and localization of neural activity, crucial for both clinical and research applications. In clinical contexts, such precision can enhance the diagnosis and monitoring of neurological conditions, where understanding the exact origins of abnormal neural activity is critical.

FEDE allowed to replicate both spatial and time-frequency features of EEG activity. Several work have attempted the replication of PSD^7,42,44^ computed from electrophysiological recordings, no one reporting the high fidelity obtained with FEDE. The replication of experimental EEG functional connectivity with personalized brain modeling has instead never been attempted before to the best of our knowledge. Several studies focused on the replication of FC matrices deduced from functional MRI (fMRI) recordings^45–49^. Modeling fMRI functional connectivity is more feasible that reproducing functional connections in EEG, as the high spatial resolution of functional MRI allows for precise localization of cortical sources and reduced volume conduction effects, which heavily affect EEG FC metrics^39^.

The introduction of a module for parameter analysis led to the identification of structural culprit of experimental EEG features. This module could be crucial for the translational use of FEDE, as it allows to reconstruct from EEG recordings key parameters related to structural brain mechanisms like synaptic transmission and noisiness of brain activity. This holds great promise for the clinical use of the model. In the case of our patient, FEDE identified as key structural parameters aberrant values of EI ratio and background noise. (both quantities are currently hypothesized to concur in determining the ASD pathophysiology^24,40^). FEDE allows to test multiple hypothesis about ASD underpinning neural mechanisms for the specific patient, which could potentially be different from one patient to another due to the heterogeneity of ASD; the model was prepared also to test hypothesis about conduction velocity and local/global connectivity impairments, however the lack of a healthy control group for comparison did not allow to test them (this being our next focus). It is also important to consider that ASD could potentially be the result of multiple concurrent impairments due to a non-typical neurodevelopment, and this can be tested using the current pipeline (see for instance concurrent combination of high background noise and high EI ratio found in the current subject); this can also be generalized to a multitude of conditions and recording techniques. FEDE offers a unique method for investigating the presence of these alterations from non-invasive recordings, nowadays unfeasible with current methods.

The choice of testing our pipeline with the creation of the first digital brain twin of a toddler patient affected by ASD also holds important implication. This complex task has never been attempted before, as both modelling a toddler brain and capturing the complexities of ASD present unique challenges yet to be addressed by current models. Working with very young subjects presents unique challenges, particularly in the acquisition and preprocessing of MRI and EEG data, affected by subject restlessness, causing several motion artefacts. By incorporating local connectivity into our models and developing a detailed representation of cortical surface activity, we have provided a novel framework for exploring the structural and functional alterations associated with ASD. The realization of a high-fidelity digital twin replica of a toddler with ASD underscores the potential of personalized precision medicine to revolutionize the diagnosis and treatment of neurodevelopmental disorders, paving the way for more effective and individualized therapeutic strategies.

Our study presents several limitations that should be addressed in future research. Firstly, the primary limitation is that our work was conducted on a single patient diagnosed with ASD. While this allowed us to demonstrate the feasibility of creating a digital brain twin in such a complex case, it limits the generalizability of our findings. The absence of a control group or additional subjects further restricts our ability to draw definitive conclusions about the broader applicability of the pipeline. A control subject or group would have provided a valuable baseline to compare the digital brain twin’s performance, enhancing the robustness of our results. This will be key in future activities. Additionally, our computational pipeline has yet to be validated across a diverse range of patients with varying conditions. The complexities of brain development and ASD are unique, and while our approach seems to be promising, it remains uncertain whether it will perform equally well in other populations, such as older children or adults with ASD, or those with other neurological conditions. Future studies with larger and more diverse cohorts are essential for validating our pipeline and assessing its broader utility in personalized medicine. Furthermore, while in principle it is possible to model several virtual versions of real-life methods of imaging and recordings, FEDE has been tested only for the replication of EEG signals.

## Methods

### Subject recruiting, ethical approval and data acquisition

The proband is a male toddler aged between 1y and 3y with a diagnosis of ASD formulated by a multi-disciplinar team according to the Diagnostic and Statistical Manual of Mental Disorders-Fifth Edition^50^ criteria, and supported the administration of the ADOS-2 Toddler Module^51^. The child’s developmental level was measured by an experienced psychologist through the Griffiths Scales of Child Development 3rd Edition^52^, and the following scores were obtained: scale A (Foundations of Learning) 99; scale B (Language and Communication) 60; scale C (Eye and Hand Coordination) 105; scale D (Personal–Social–Emotional) 88; scale E (Gross Motor) 105. Moreover, the Childhood Autism Rating Scale second edition (CARS-2)^53^ was administered by a qualified evaluator with clinical experience in ASD, and resulted in an overall score of 36, which indicates a level of moderate autism. He underwent both neuroimaging and neurophysiological investigations for clinical purposes at a tertiary care university hospital, the IRCCS Fondazione Stella Maris (Pisa, Italy), in order to exclude brain anomalies. The electroencephalogram (EEG) (Micromed, Mogliano Veneto, Italy) study was performed according to the 10-20 International System. Brain magnetic resonance imaging (MRI) was obtained with a GE 3T scanner (Signa HDx, GE-Medical-Systems, Milwaukee, United States) using a protocol optimized for neurodevelopmental disorders which included three-dimensional (3D) T1-weighted (T1W) fast-spoiled gradient echo, 3D T2 fluid-attenuated inversion recovery, two-dimensional (2D) T2-weighted (T2W) fast spin echo, and diffusion weighted imaging. This study was conducted in accordance with the Declaration of Helsinki and approved by the Regional Ethical Committee of Meyer Hospital (Florence, Italy), number 131/2024. Informed consent for the study was provided by the patient’s legal guardians.

### Pipeline Overview

T1-weighted (T1-w), T2-weighted (T2-w) and Diffusion-Weighted Imaging (DWI) acquisitions from a single pediatric subject (aged between 1y and 3y), provided by IRCSS Stella Maris, were processed to create the digital brain model. Due to the clinical data not meeting the mandatory requirements of the ‘TVB Image Processing Pipeline’ — specifically lacking B0 Reverse Phase Encoding^12^ (RPE), having only one non-zero B-value, and missing FLASH acquisition^14^ — a tailored pipeline was developed. This customized pipeline integrates multiple tools and software packages to achieve the necessary preprocessing and analysis:

- **FSL and ANTs**: Employed for preprocessing steps such as denoising, unwarping, removal of eddy currents, bias correction, and brain mask generation.
- **MRtrix3**: Utilized for tractography, constructing the connectome, computing structural connectivity (SC) weights, mean tract lengths and conduction velocity.
- **FreeSurfer**: Used for anatomical reconstruction, cortical mesh creation, and segmentation/parcellation of the brain.
- **SimNIBS**: Applied to create the Head Model and the EEG Forward Leadfield Matrix
- **MNE (MNE-Python)**: Implemented to generate inputs for **The Virtual Brain** (TVB).

This integrated approach ensures that the subject-specific data, despite its limitations, can be effectively processed and used as inputs for advanced brain modeling and analysis within TVB.

### MRI acquisition

The subject data were acquired in a clinical setting at IRCSS Stella Maris in Pisa (Italy). MRI was acquired with a 3T General Electric scanner; T1 with voxel size of 1.0mm by 0.4297 mm by 0.4297 mm, TR=2471.42ms, TE=3.824ms; T2 with voxel size of 0.799805mm by 0.5 mm by 0.5 mm;DTI with voxel size of 0.9375mm by 0.9375mm by 5.0mm 30 gradient encoding directions and B=1000s/mm2 TR=3100ms, TE=59.6ms.

### MRI Preprocessing

The preprocessing phase involved several critical steps to prepare the imaging data for further analysis^9,54^. Initially, the DWI data underwent denoising to remove noise artifacts and removal of Gibbs’ ringing artifacts, followed by unwarping to correct for geometric distortions^55^ Concerning this latter, since clinical data were lacking the Reverse Phase Encoding, a synthetic RPE was created using SynB0-DISCO^12,56^.

Eddy current-induced distortions were also corrected^57^, and bias correction was applied to mitigate intensity inhomogeneities^9^. A brain mask was generated to isolate the brain from non-brain tissues. Preprocessing steps were performed using FSL^58^ and ANTs^59^ softwares. A possible different segmentation of subcortical structures (by using Freesurfer Infants^60^) was also tested with the current subject, with no relevant differences in segmentation results. Images were subsequently preprocessed following the preprocessing pipeline outlined in Andy’s Brain Book (Jahn, 2022. doi:10.5281/zenodo.5879293).

### Constrained Spherical Deconvolution

To characterize the white matter structure, a constrained spherical deconvolution (CSD) was performed^10^. This process included estimating the white matter, cerebrospinal fluid and gray matter response functions for the subject, generating fiber orientation densities^61^ (FODs) and normalizing them. Due to the limited diffusion data available (only one gradient direction with B=1000s/mm²), the response functions were generated using MRtrix3Tissue (https://3Tissue.github.io), a fork of MRtrix3^62^l.

### Tissue Boundaries and Coregistration

Following deconvolution, tissue boundaries were created, and the diffusion images were co-registered with the anatomical T1-weighted images^63^. The boundary interface between gray matter and white matter was identified and used as a seed region for streamline generation through Anatomically Constrained Tractography^13^ (ACT). This step was performed to ensure that the tractography results were anatomically accurate, providing a robust foundation for subsequent connectivity analysis and brain modeling.

### Probabilistic Tractography

Probabilistic tractography^64^ was performed using Constrained Spherical Deconvolution^10^ coupled with Anatomically Constrained Tractography^13^ and Spherical-deconvolution Informed Filtering of Tractograms^65^ (SIFT2). Dynamic seeding was also employed to ensure robust streamline generation. The maximum length of fibers was set to 250 mm, following the guidelines of the TVB Image Processing Pipeline^66^. We chose a number of streamlines proportional to the numerosity of regions considered in the parcellation (1000 streamlines for each pair of regions). Specifically, this resulted in a number of 71.631 million streamlines for the adopted HCPMMP1^35^ atlas.

### Anatomical Reconstruction

Anatomical reconstruction was conducted using FreeSurfer’s ‘recon-all’ command, which performed cortical mesh creation and segmentation/parcellation using HCPMMP1 atlas. The ‘-autorecon1’ command executed the initial five steps, with tailored inputs to improve subsequent skull extraction. Brainmask was checked and small manual adjustments performed. After the ‘- autorecon2’ the white matter mask was checked and slightly edited followed by autorecon2-wm; at last, the ‘-autorecon3’ command completed the remaining steps of the anatomical reconstruction process.

### Structural Connectivity Weights, Mean Tract Length

We employed MRtrix3^62^ to generate the connectome and calculate structural connectivity (SC) weights, intended as weighted (since SIFT2 was used) number of streamlines connecting two regions, and mean tract length

### Myelin volume fraction, g-ratio and conduction velocity

We conducted a comprehensive preprocessing and analysis of MRI data to derive conduction and myelinization properties^67^. It has been followed the methodology from Newman et al.^16^. Initially, the T2-w was co-registered to T1-w.

Brain extraction was performed on the T1-w and T2-w images using the bet function from FSL with specific fractional intensity and gradient threshold settings to generate brain-only images and their masks. The intensity values of these images were then standardized. Minimum and maximum intensity values of T1 and T2 images were computed using fslstats, and the images were rescaled to a 0-100 range via fslmaths.

Subsequently, a T1/T2 ratio map was generated. We then extracted fixels (“fiber population within a voxel”) from the normalized wmfod using MRtrix3 fod2fixel and computed the sum of the Apparent Fiber Density (AFD) using fixel2voxel.

Coregistration and regridding was achieved by aligning the fixel-derived apparent fiber density^36^ (AFD) sum map with the T1/T2 map using the flirt command, followed by transformation matrix conversion (transformconvert) and regridding (mrtransform, mrgrid).

Calculation of the myelin volume fraction^11^ (MVF) and axon volume fraction (AVF), are derived by the method described by Mohammadi and Callaghan^68^, computing MVF/AVF ratios and generating a voxel-wise g-ratio map using fslmaths. The equation for the g-ratio is:

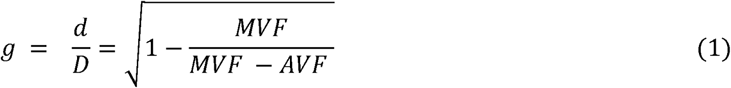

where (d) is the axon diameter and (D) is the external myelinated fiber diameter.

This g-ratio map was subsequently upper-thresholded. We further computed the voxel-aggregated conduction velocity (CV) based on the derived g-ratio and AVF using the model by Berman, Filo, and Mezer^15^, according to the equation:

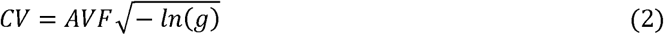

The CV map was then coregistered with diffusion-weighted imaging (DWI) to align with streamlines from tractography.

Finally, the CV map was sampled along the streamlines of tractography to extract mean velocity per streamline (*tcksample*) and from this latter the CV matrix was calculated (*tck2connectome*).

### Cortex Surface Processing

In order to reduce computational time, the cortical mesh was downsampled from FreeSurfer’s original fsaverage(7) with 327680 triangles to the fsaverage5, which has 20,484 vertices and 40,960 triangles (for comparison, the TVB default cortical mesh^2^ uses 16,384 vertices and 32,760 triangles).

### Reconstruction of Head Model and Calculation of anatomically-accurate EEG Forward Leadfield Matrix

The replication of EEG signals requires projecting neural activity from the high-density cortical mesh to the electrodes on the scalp surface, computing a mathematical quantity called lead-field matrix. The lead-field matrix is a *n_vertices_×n_electrodes_* matrix, whose entries encode how the activity of each vertex in the cortical mesh can influence the signal in each EEG electrode. We implemented a pipeline to determine patient-specific lead-field matrix based on patient anatomy. Anatomical tissues between cortex and scalp electrodes were reconstructed with the SIMNIBS software (see Methods).

In order to calculate the EEG Forward Problem Leadfield Matrix, it has been first reconstructed the Head of the subject with 12 tissues using SimNIBS4.1.0: the T1-w and coregistered/regridded T2-w were used as input to *charm* command together with the Freesurfer anatomical reconstruction folder and the initial transformations (the SimNIBS template was coregistered to the T1-w using Freesurfer to find the Transformation matrix used to initialize the affine registration of the SimNIBS template to the subject MR), then also the DWI image was used with the *dwi2cond* command (based on FSL *dtifit*), in order to calculate the tensors necessary to compute the anisotropic properties of conductivity for GM and WM (see Supplementary Figure S5) using the “Volume Normalized” algorithm as shown by Opitz et al^17^

It has been chosen an EEG cap from Neuroelectrics following the 10-10 system^69^ and with 64 channels (see Figure 1) using electrodes with circular shape (10mm diameter and 4mm of saline gel). This latter, together with the Head Model, the conducitivities shown in Supplementary Figure S2, and the calculated anisotropic properties, allowed to calculate the EEG Leadfield Matrix using the *TDCSLEADFIELD* algorithm.

### TVB Format Conversion

The next step involved the use of MNE to rearrange the cortical mesh and prepare inputs for The Virtual Brain (TVB). A Python script, derived from ‘convert2TVB_format.py’^66^, was employed to achieve this.

The script generates for each vertex the pertaining region, hemisphere, and assigns a flag indicating the vertex to be either in a cortical or in a non-cortical area. We computed a mid-thickness cortical surface by averaging the white and pial surfaces (this in order to match one to one the SimNIBS cortex), with a dummy region for non-cortical areas^70^. Also it writes the ‘Leadfield’ Matrix in TVB format, convert the vertices coordinates from Freesurfer system (ras-tkr) to image coordinates (Subject World) and write the cortex file in zipped TVB format (normals, triangles, vertices), then converts electrodes coordinates (again from FS ras-tkr to image coordinates “Subject World”) and write the EEG sensors locations in TVB format, write in zipped TVB format the SC weight, Mean Tract Length, Mean Velocity, Region Centres, Average Orientation (mean normal of vertices for a triangle).

As last step, the local connectivity matrix of the cortical surface, providing the weight of the lateral connectivity between cortical columns (assumed as instantaneous), was computed using a gaussian distribution (where amplitude and sigma were chosen after the parameter fitting) applied to the geodesic distance on the mid-thickness cortex. Also, conversion to h5 format was performed in order to use the visualizer of TVB.

To compare the anatomically-accurate forward solution with current standard methodology, we created the BEM model of the patient’s with Brainstorm, importing the result of SimNIBS charm and then considering only the interfaces between brain tissue, skull and scalp. Default conductivity values (measured as conductance over length, S/m) were assigned to different volumes: 0.465 S/m for brain compartment, 0.008 S/m for skull compartment, and 0.465 S/m for scalp compartment^71^ (this because for BEM method it is the ratio between compartments driving the results, which is normally 1:1/80:1,^18^ however it was reduced in order to account for the young age of the subject). The leadfield matrix was computed with OpenMEEG^43^. BEM surfaces and electrodes coordinates were converted from the FreeSurfer system (ras-tkr) to the image coordinate system (ras-scanner), and the lead field matrix was converted into TVB format with the same procedure adopted for the anatomically-accurate one.

### Importing Input Files into The Virtual Brain and Creating Brain Simulation

SC data, conduction velocity, Region mapping, cortical vertices, electrode positions and local connectivity were imported in TVB to perform the simulations (h5 were created for import and visualization purposes in TVB GUI). Activity on cortical vertices was simulated by using the Jansen-Rit model^37^. The Jansen-Rit cortical was chosen for its capacity of replicating EEG signals^37^. Subcortical structures were minimally segmented and modeled using the same Jansen-Rit model as the cortex. The simulation of cortical surface activity allows the modeling and analysis of both local and global connectivity^2^, as both are reported to be aberrant in ASD^26,30,72,73^. An anatomically-accurate forward solution was used to compute virtual EEG from simulated activity^74^. As already mentioned, the Local connectivity was computed from geodesic brain distance by using a gaussian kernel, whose extension and strength was determined by parameter fitting.

### Parameter Fitting with Experimental EEG Recording

To tune model parameters using the experimental EEG recording, a hierarchical grid search procedure was implemented (see Results). Parameters varied including white matter speed, SC weights scaling, sigmoidal Jansen-Rit coupling, local connectivity, and Jansen-Rit model parameters. For each combination of parameters, a simulated EEG was generated, with a total of 1480 simulations.

Simulated signals were fitted to experimental recordings, based on a loss function that computes the difference in functional connectivity and power spectral densities. The two loss functions are:

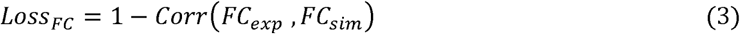

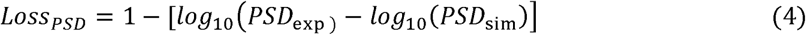

Where *Corr* is the standard Pearson correlation. In the _PSD_ function, the PSD were truncated at 20 Hz (this in order to improve the efficacy of the loss function in locating the best fits). The best match between simulated and actual EEG was computed using these metrics, thus identifying the optimal parameter values.

### Experimental EEG acquisition and pre-processing

EEG data were acquired using the Micromed System Plus Evolution with a sampling rate of 256 Hz, utilizing a 21-electrode sensor system adhering to the standard 10-20 layout. The recorded signals were imported into EEGLAB^75^ for further processing. Initially, the resting state epoch was extracted from the continuous EEG recording to capture the subject’s brain activity in a relaxed state without any task-specific stimuli. The EEG data were then filtered in the 1-40 Hz range using a Hamming windowed sinc filter, following an initial high-pass filtering at 1 Hz to remove slow drifts and baseline wander, ensuring the relevant frequency components were retained for analysis.

To enhance the quality of the EEG signals, the *cleanrawdata* plugin in EEGLAB was used to identify and remove bad channels and segments. Channels that were silent or flat for more than 5 seconds, exhibited a large amount of noise based on their standard deviation (with a rejection threshold of 4 standard deviations), or had low-frequency signals poorly correlated with nearby channels (using a threshold value of 0.8) were removed. This resulted in a final number of 16 channels selected for further experimental analysis. Additionally, bad portions of the data series were rejected using the Artifact Subspace Reconstruction^76^ algorithm for regions exceeding 20 times the standard deviation of the calibration data, and further rejection was based on how many channels within a specified time range exceeded a standard deviation threshold, with a maximum out-of-bound channel percentage set at 25%.

The remaining data were re-referenced using the average reference method^77^, averaging the signals across all electrodes and subtracting this mean from each electrode. Independent Component Analysis (ICA) was then performed using the infomax algorithm to decompose the EEG signals into independent components, which were classified using the ICLabel plugin^78^. Components identified as artifacts (e.g., eye blinks, muscle activity, or electrical noise) were removed from the dataset, ensuring that only clean EEG signals were retained for further analysis. This preprocessing ensured a robust foundation for accurate characterization of neural activity and the validity of subsequent analyses.

### EEG features: power spectral distribution

We computed the power spectral distribution (PSD) using Welch’s method, dividing the signal into overlapping segments, applying windowing, and taking the discrete Fourier transform to obtain power spectrum estimates. These estimates were averaged to reduce variance, producing a binned array of power spectral densities for each frequency.

Oscillations in neural data are embedded within aperiodic activity, typically following a 1/f distribution. Traditionally, this aperiodic component was disregarded or treated as noise. However, variations in aperiodic activity are now recognized as potential biological indicators for disease, aging, and development. Therefore, we parametrized PSDs into periodic and aperiodic components using the FOOOF (fitting oscillations and one-over f) algorithm^79^.

FOOOF models the PSD as a combination of 1/f activity *L* and *N* periodic components *G_n_*:

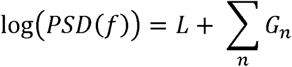

With periodic components *G_n_* modeled with Gaussians and the aperiodic component *L* is modeled as:

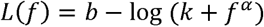

where *b* is the broadband offset, *α* is the exponent, and *k* is the knee parameter.

FOOOF involves initial fitting of the aperiodic component, detrending the spectrum, detecting and fitting periodic components iteratively, and combining these fits to create the model, computing goodness-of-fit scores. If the knee parameter *k=0*, aperiodic exponents are more comparable and interpretable, serving as potential biomarkers.

### EEG features: functional connectivity

Functional connectivity was computed with standard Pearson correlation, considering only significant (p<0.05, Bonferroni correction) values. The choice of the Pearson correlation metric was motivated by the limited number of electrodes^39^. Furthermore, we adopted this metric to prove that the FEDE pipeline is able of replicating also FC measures heavily affected by volume conduction^80^.

## Data Availability

Code and patient data can be transmitted upon reasonable request to

## Acknowledments

We thank Adriana di Martino, Ting Xu, Emiliano Ricciardi, Benjamin T Newman, Jan Paul Triebkorn, Jurek Timo Hofsähs and Marco Pagani for fruitful discussions on different aspects of the pipeline.

## Author Contributions

MF conceived and performed the simulations and analysis and ideated the MRI preprocessing pipeline; LGA conceived and performed the simulations and analysis and wrote the paper; LM preprocessed experimental EEG data and performed the simulation and analysis; JC performed the simulation and analysis; EB and SC collected experimental MRI and EEG data; AR conceived the simulations and analysis; AAV conceived and performed the simulations and analysis; AM conceived the simulations and analysis and obtained the fundings that financed the project.

