## Supplementary Materials for "Reconstructing whole-brain structure and dynamics using imaging data and personalized modeling"

**Table S1.**

| **P****arameter** | **Description** | **Measure unit** | **Value** |
| --- | --- | --- | --- |
| *A* | Maximum Excitatory | mV | 3.50 |
|  | post-synaptic potential |  |  |
| *B* | Maximum Inhibitory | mV | 12.0 |
|  | post-synaptic potential |  |  |
| *τe* | Excitatory time constant | ms | 8.3 |
| *τi* | Inhibitory time constant | ms | 50 |
| *v*0 | Voltage threshold for which a | *mV* | 6.0 |
|  | 50 % firing rate is achieved |  |  |
| *νmax* | Maximum firing rate of | s−1 | 0.0025 |
|  | neural population |  |  |
| *r* | Steepness of sigmoidal | mV−1 | 0.56 |
|  | transfer function |  |  |
| *C*1 | Average probability of synaptic contacts | pure number | 1.2 |
|  | in the feedback excitatory loop |  |  |
| *C*2 | Average probability of synaptic contacts | pure number | 0.8 |
|  | in the slow feedback excitatory loop |  |  |
| *C*3 | Average probability of synaptic contacts | pure number | 0.25 |
|  | in the feedback inhibitory loop |  |  |
| *C*4 | Average probability of synaptic contacts | pure number | 0.3 |
|  | in the slow feedback inhibitory loop |  |  |
| *η* | Mean input noise | s−1 | 0.088 |
| *µ* | Mean input firing rate | s−1 | 0.12 |
| *J* | Average internal connections | pure number | 150 |
| WM speed | Multiplicative constant to the conduction velocity matrix | pure number | 30 |
| WM coupling (a) | Multiplicative constant to the structural connectivity matrix (normalized with nr of generated streamlines during tractography) | pure number | 1.86*10 ^-5^ |
| WM coupling (cmin) | Minimum of the sigmoid function | pure number | 0.0 |
| WM coupling (cmax) | Maximum of the sigmoid function | pure number | 0.005 |
| WM coupling midpoint | Midpoint of the linear portion of the sigmoid |  | 3.12 |
| WM coupling (r) | Steepness of the sigmoidal transformation | pure number | 0.28 |
| LC strength | Strength of local connections | pure number | 0.1 |
| LC sigma | Area of influence of local connections | pure number | 0.5 |

**Table S1: Optimal combinations of model parameters:** Parameters of the Jansen-Rit neural mass model are reported first, and separated from the parameters of the brain mode by a horizontal line

**Table S2.**

|  | **KS test** | **p-value** |
| --- | --- | --- |
| **Delta** | 0.53 | 0.10 |
| **Theta** | 0.33 | 0.38 |
| **Alpha** | 0.46 | 0.30 |
| **Beta+Gamma** | 0.46 | 0.30 |

**Table S2: Statistical analysis of differences in PSD relative power between simulated and experimental EEG channels.** Kolmogorov-Smironv test was used, highlighting no statistical differences between the two distributions.

**Figure S1.**

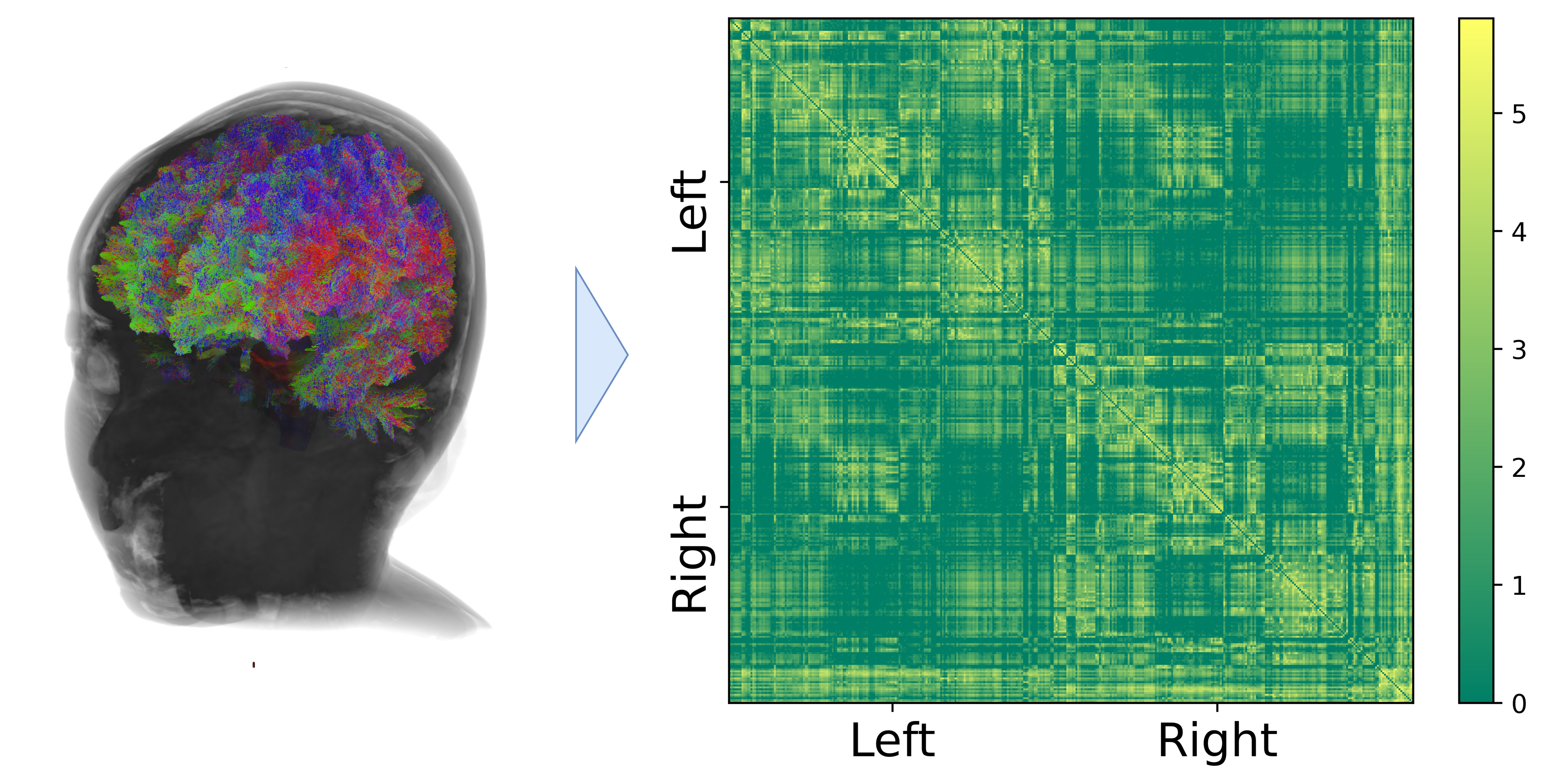

**Figure S1: Structural connectivity matrix.** Structural Connectivity weights between brain regions, reported in logarithmic scale (log_10_(1+weight)). Structural connectivity matrix is derived from tractography analysis combined with gray matter parcellations. Connections are arranged according to hemispheres subdivisions, while subcortical regions are reported in the bottom and right part of the matrix.

**Figure S2.**

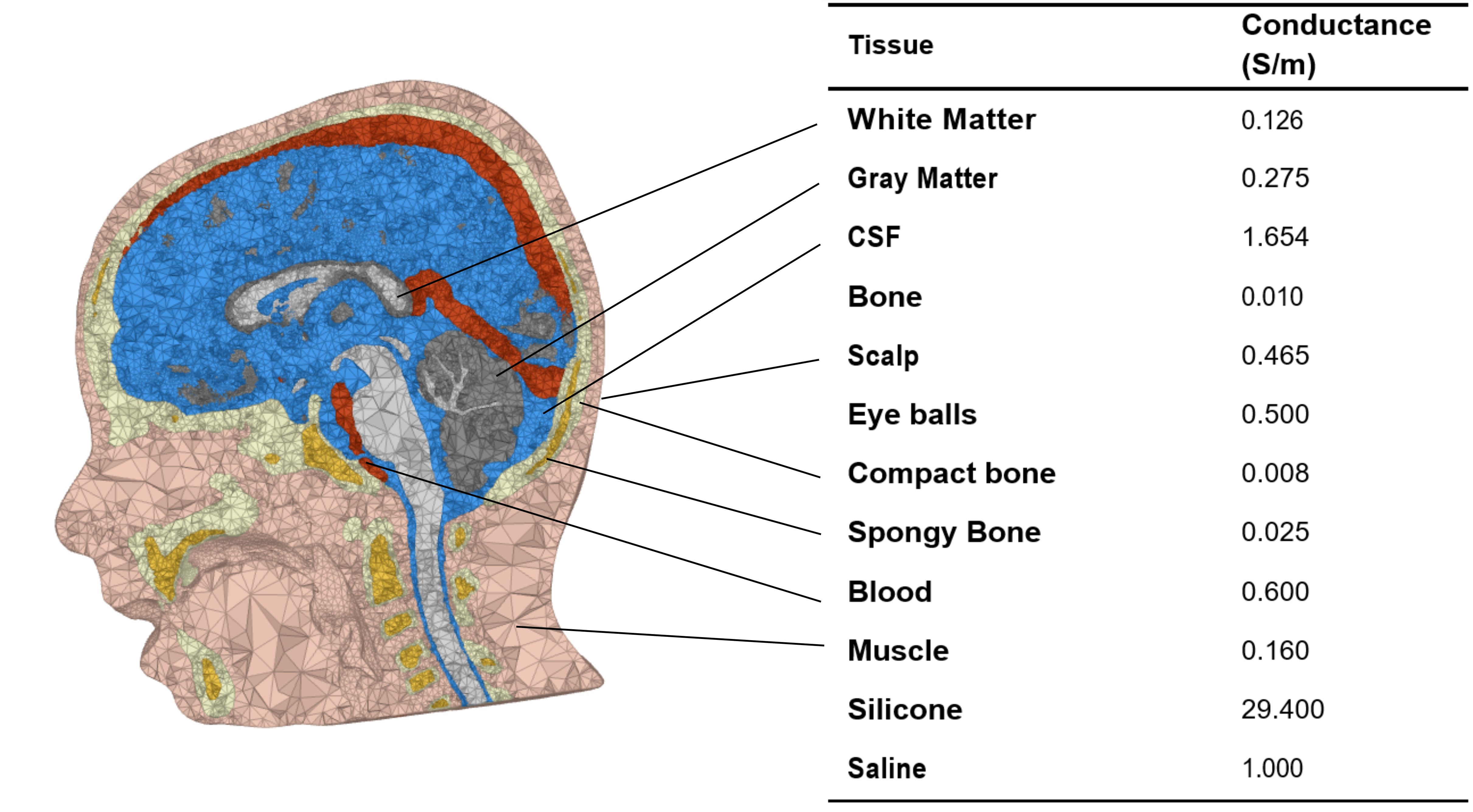

**Figure S2: 3D FEM model of patients’ head with tissue and electrodes conductances.** Values can be consulted in <https://simnibs.github.io/simnibs/build/html/documentation/conductivity.html>, alongside references from which the single conductance values are deduced.

**Figure S3.**

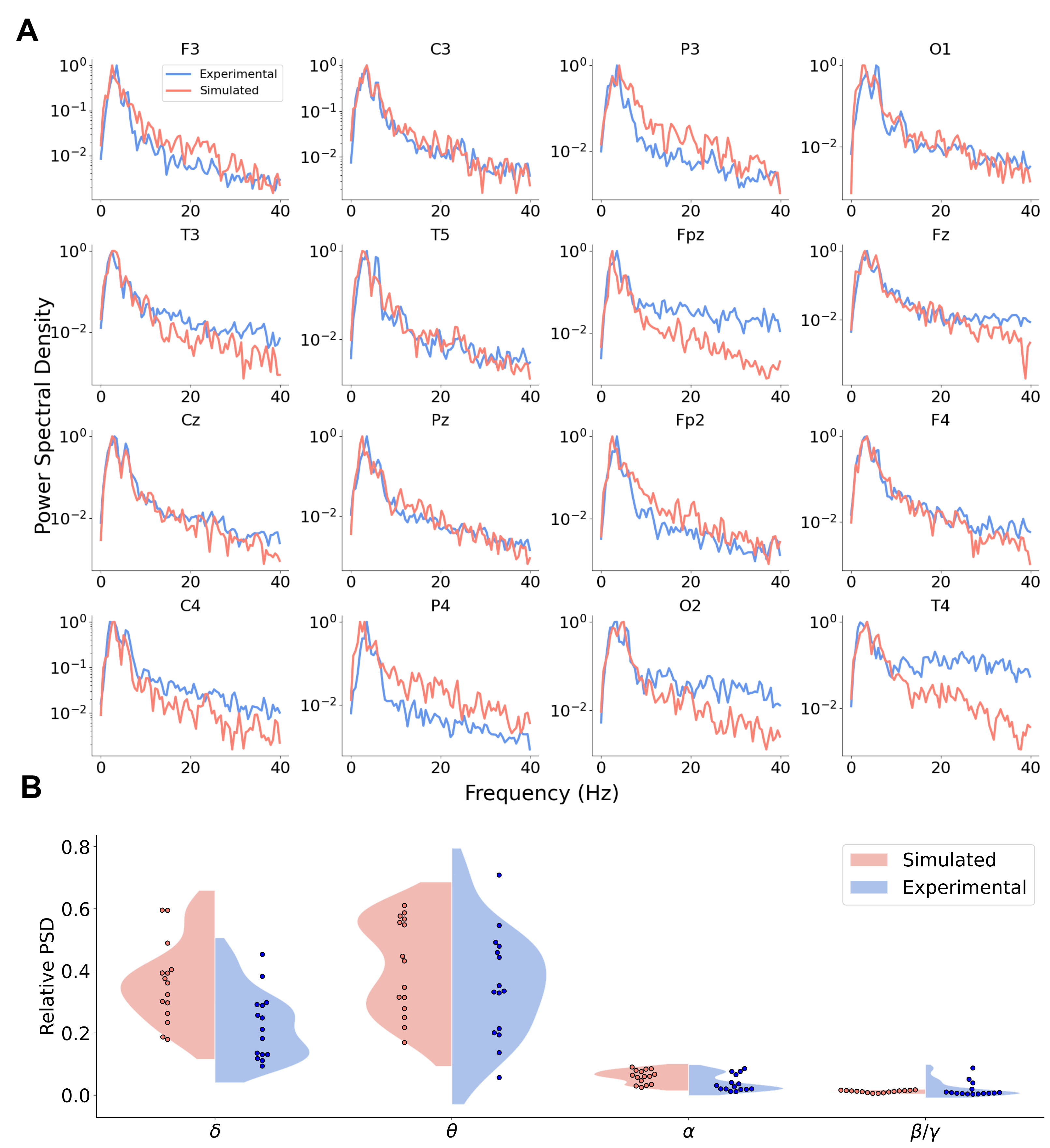

**Figure S3: Channel by channel comparison of experimental and simulated PSDs. (A):** Single channels PSD from experimental and simulated signals. **(B):** Violin plot of relative EEG band power of channels. Distribution did not present statistically relevant differences between experimental and simulated signals.

**Figure S4.**

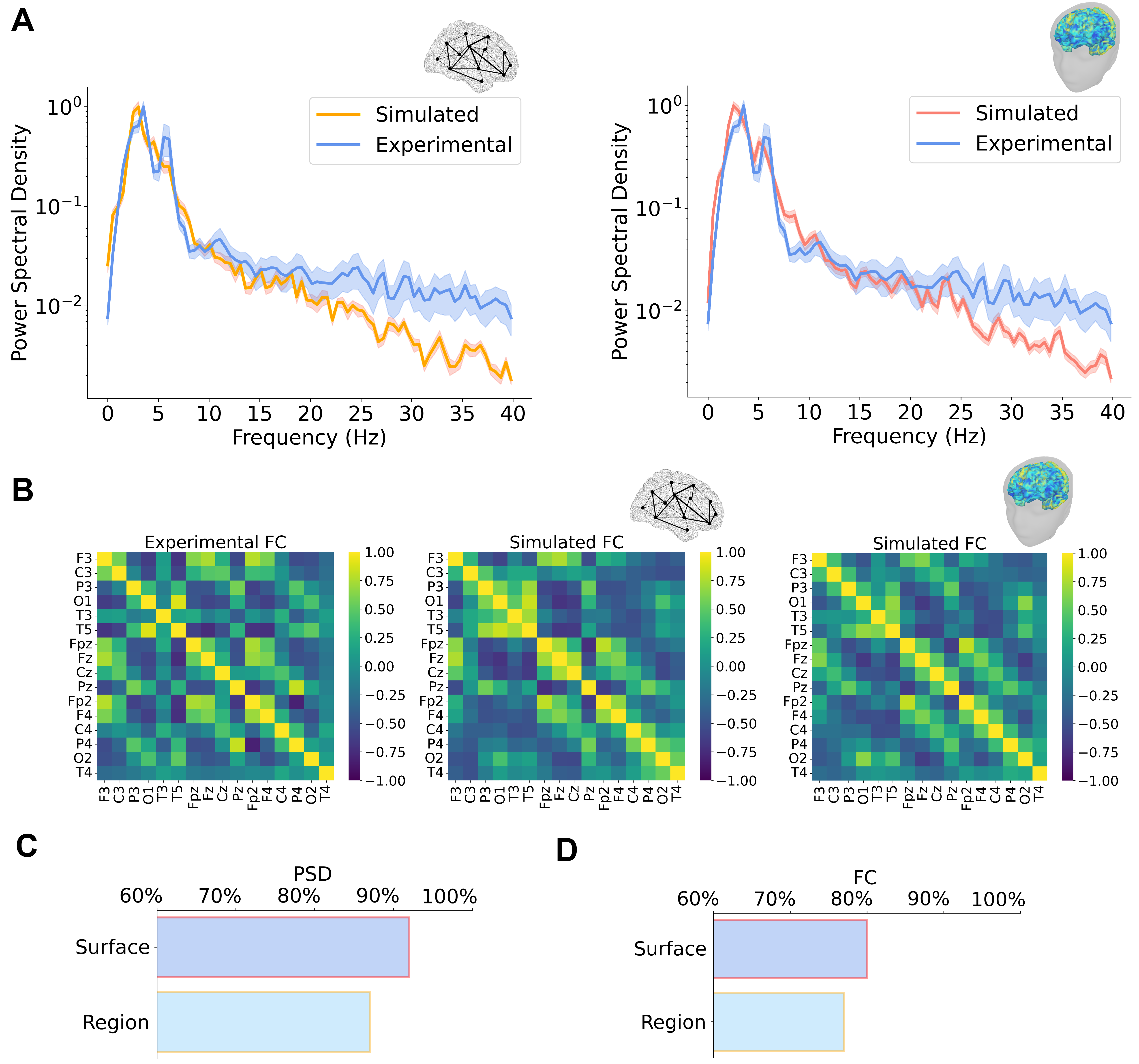

**Figure S4: Comparison between simulated PSDs and FCs with region and surface-based simulation.** Model parameters selected through parameter exploration were utilized in a region-based simulation, using as scaffold the 379-regions HCPMMP1 atlas employed for gray matter parcellation. Results were compared with both the experimental values and with surface-based simulations. **(A):** PSD computed from region-based simulations fails to capture finer details of the experimental PSD, such as the second peak in low-alpha band. **(B):** FC matrices computed from region-based simulations show a more stereotypical differentiations between highly and low functionally connected electrode. **(C):** r-regression coefficient between experimental and simulated PSD is higher for surface-based analysis. **(D):** Similarly, r-coefficient between experimental and simulated FC is higher for surface-based analysis.

**Figure S5.**

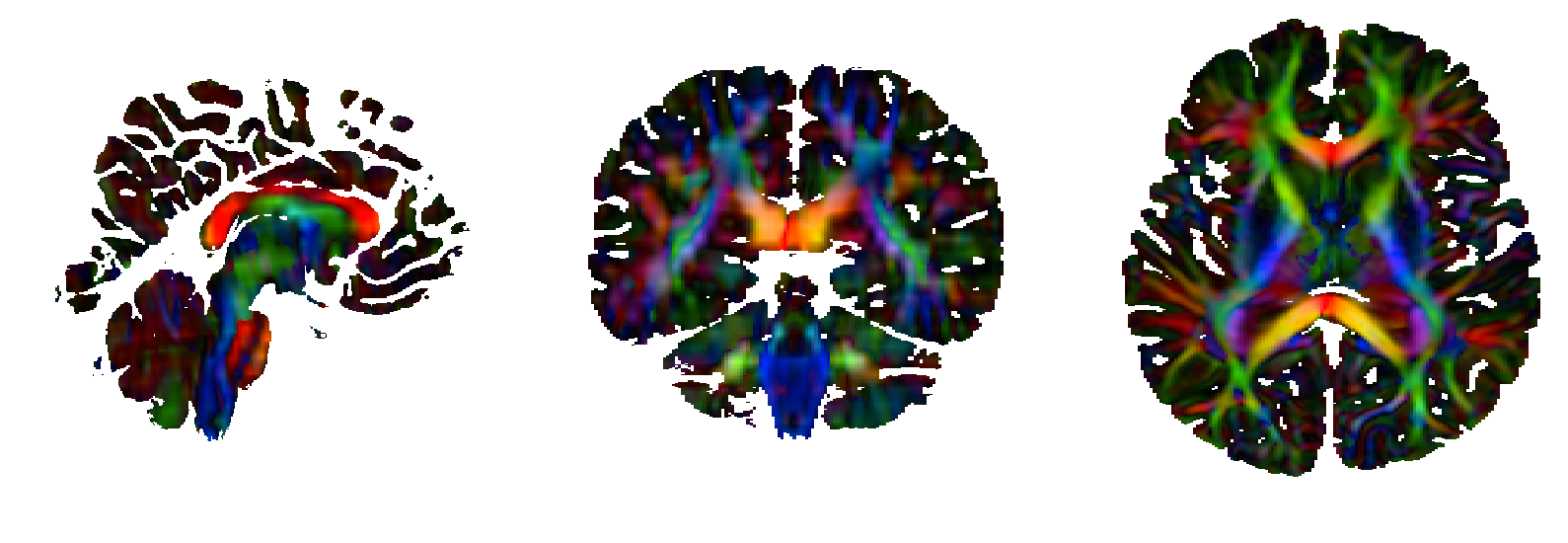

**Figure S5: Determination of structural tissue anisotropy.** The DWI image was used as input to the *dwi2cond* command (which is based on FSL *dtifit* algorithm), in order to calculate the tensors necessary to calculate the anisotropic properties of conductivity for GM and WM using the “Volume Normalized” algorithm (see main text).
